# Rural Disparities in Early Childhood Well Child Visit Attendance

**DOI:** 10.1101/2020.11.10.20229179

**Authors:** Pamela B. DeGuzman, Guoping Huang, Genevieve Lyons, Joseph Snitzer, Jessica Keim-Malpass

## Abstract

**Purpose:** Children should attend well child visits (WCVs) during early childhood so that developmental disorders may be identified as early as possible, and if indicated treatment can begin. The aim of this research was to determine if rurality impacts access to WCV during early childhood, and if altering rurality measurement methods impacts outcomes.

**Design and Methods:** We utilized a longitudinal correlational design with early childhood data gathered from the Virginia All Payer Claims Database, which contains claims data from Medicaid and the majority of Virginia commercial insurance payers (n=6349). WCV attendance was evaluated against three rurality metrics: a traditional metric using Rural-Urban Commuting Area codes, a developed land variable, and a distance to care variable, at a zip code level.

**Results:** Two of the rurality methods revealed that rural children attend fewer WCVs than their urban counterparts, (67% vs. 50% respectively, using a traditional metric; and a 0.035 increase in WCV attendance for every percent increase in developed land). Differences were attenuated by insurance payer; children with Medicaid attend fewer WCVs than those with private insurance.

**Conclusions:** Young children in rural Virginia attend fewer WCVs than their non-rural counterparts, placing them at higher risk for missing timely developmental disorder screenings. The coronavirus disease pandemic has been associated with an abrupt and significant reduction in vaccination rates, which likely indicates fewer WCVs and concomitant developmental screenings. Pediatric nurses should encourage families of young children to develop a plan for continued WCVs, so that early identification of developmental disorders can be achieved.

---

Well child visits (WCVs) during early childhood (ages 1-4) are an important component of pediatric preventive health care. Screening for developmental disorders is predominantly performed during early childhood, at the 18-, 24- and 30-month visits, supplementing the continuation of immunizations and growth evaluations that were initiated during infancy (American Academy of Pediatrics Committee on Practice and Ambulatory Medicine & American Academy of Peditrics; Bright Futures, 2017). By 18 months of age, delays in communication and language development can be detected and are amendable to intervention; by 30 months, cognitive delays are also both detectable and addressable (Duby et al., 2006).

Developmental screening during early childhood is particularly important to promote identification of ASD. ASD can be reliably diagnosed by two years of age (Kleinman et al., 2008; Lord et al., 2006). Earlier ASD intervention is critical, as it has been shown to considerably improve treatment outcomes (MacDonald et al., 2014; Mazurek et al., 2014). Rural children have fewer opportunities than their urban counterparts to be diagnosed with ASD. Although one in six young children living in a rural area has a mental, behavioral or developmental disorder diagnosis (Robinson et al., 2017), rural children with special healthcare needs are less likely than their urban counterparts to be seen by a pediatrician (Skinner & Slifkin, 2007). Missing WCVs during early childhood can further impact the ability for rural children with a developmental delay to receive appropriate care, because many rural areas already have limited access to diagnostic specialists such as developmental pediatricians (DeGuzman, Altrui, et al., 2017; Rosenberg et al., 2011).

Although some disparities in pediatric preventative health visits have been documented, rural disparities in WCV attendance are largely unexplored. A national study reporting data from 2007-2008 found that overall WCV attendance rates to be close to 60%, with lower rates of attendance driven by lower family income, less parental education, minority race or ethnicity of the child, and lack of insurance coverage (Abdus & Selden, 2013). Access to children’s insurance has improved in many U.S. states with the 2010 passage of the Affordable Care Act, but insurance coverage itself is unlikely to completely resolve disparities in WCV attendance when other contextual social determinants of health that drive limited access remain. In fact, one study that only included insured children found that 60% of 0-2 year-olds and 40% of 3–5 year-olds had fewer WCVs than recommended (Goedken et al., 2014). Despite the importance of WCVs and the higher prevalence of developmental disorders in rural children, no studies were found that have evaluated the differences in children’s attendance at WCVs between those living in rural and urban areas. Understanding the impact of rurality on WCV attendance can help policy makers and practitioners better target programs to ensure adequate child health access.

Measuring rural health disparities is challenging due to the lack of a singular definition of “rural” in the disparities literature, none of which may be adequate to capture known drivers of rural health disparities (Coburn et al., 2007). Accordingly, research has shown that application of different definitions may produce different results (Meilleur et al., 2013; Stern et al., 2010). Several classification codes are available only at a county level, limiting the ability of researchers to understand the impact of more granular variations in factors that drive disparities such as distance to care, or landscape-related hazardous roads which can vary widely in large area rural counties (DeGuzman, Cohn, et al., 2017; Pesut et al., 2010). For example, the National Center for Health Statistics (NCHS) Urban–Rural Classification Scheme for Counties was developed specifically to differentiate geographic health disparities, but is available only at a county level (Ingram & Franco, 2014). In states that are characterized by a range of settings within counties, using a county-level approach is problematic. Virginia is a largely rural state, with densely populated metropolitan areas in northern Virginia, Richmond, and the Tidewater (Virginia Beach/Norfolk) area. Beyond its urban centers, the state quickly transitions to rural countryside, forest, or farmland interspersed with smaller cities. An example of the limitations of county-level classification is highlighted in Nelson County, Virginia. Nelson County is located directly south of, and a 30-minute drive to Albemarle County, where the University of Virginia and small city of Charlottesville are located. The county is classified in the middle of the NCHS classification range as ‘small metropolitan,’ despite the area’s mountainous terrain and low population density. Across its nearly 500 square miles, there are multiple pockets of residents with high poverty and low education levels. Although there are only 12 Census block groups (CBGs) in Nelson County, the 2017 American Community Survey reported wide disparities among the CBGs’ median incomes, ranging from $16,906 -$87,132, and percent of CBG residents stopping education at the high school level or earlier ranges from 15%-76% (*U*.*S. Census Bureau’s American Community Survey 2013-2017 5-Year Estimates, Tables B28001, B28002*, n.d.). Using county-level data blurs the distinctions between highly diversified areas across this large land area.

Rural Urban Commuting Area (RUCA) codes are a common method used to distinguish rural from urban in disparities research and can be applied to Census tracts and zip codes. However, RUCA was developed with population and commuting data, not with a healthcare lens and may overlook important healthcare contextual variables such as broadband access (DeGuzman et al., 2020). Thus, using RUCA for a more granular approach may also be problematic. Two of the three Census tracts in Nelson County are classified as “urban” despite fewer than 25% of these households having access to broadband internet, a known driver of healthcare access in rural areas (Greenberg et al., 2018). If we want to identify and address rural disparities for Virginia’s children and other rural populations experiencing geographic-related health disparities, more meaningful approaches are needed. The aim of this research was to determine if rurality impacts early childhood WCV attendance in Virginia, and to determine what differential information may be gleaned by altering the way rurality is measured.

## Methods

We employed a longitudinal correlational design to explore the study aims. A secondary data set was obtained from the Virginia All Payer Claims Database (APCD). The APCD includes paid claims from approximately 70% of Virginia commercial health insurance companies, and all claims paid through Virginia Medicare and Medicaid. Because the APCD includes insurance claims, no data about self-pay visits are included (Virginia Health Information, 2018). Our data set included all aspects of claims made including for each claim, the date of the claim, all diagnosis codes, all current procedure terminology (CPT) codes, the insurance type, and demographic information (gender, race, and zip code). The Virginia APCD was started in 2011; thus, inclusion criteria for our dataset was all children born in 2011 who had both an insurance claim for the 12 month and *pre-kindergarten* WCVs, which is required for the child to enter kindergarten. The pre-kindergarten WCV was defined as a minimum of one WCV between the ages of 4 and 6. These inclusion criteria were chosen to ensure that children who moved out of state during the early childhood years did not have missing WCVs inappropriately classified as non-attended.

### Variables

WCV attendance rates were developed from the APCD data set. WCVs were identified in the dataset as either ICD-10 codes (Z00.121, Z00.129, or V202) or CPT codes (99392, 99393, 99382 or 99383). The American Academy of Pediatrics *Periodicity Schedule* recommends WCVs during early childhood at 12 months, 15 months, 18 months, 24 months, 30 months, 3 years and 4 years (American Academy of Pediatrics Committee on Practice and Ambulatory Medicine & American Academy of Peditrics; Bright Futures, 2017). For each child, classification of WCV visits into these categories was determined by calculating the difference in time between the child’s birth CPT and the WCV code to approximate age, and then determining which of the visits (12-month, 15-month etc.) the age of the child was closest.

### Rural Classification

In order to evaluate the impact of rurality using different measurement approaches, we identified three separate ways to measure rurality. All three measures were developed using the child’s zip code, which was available in the APCD dataset. For each zip code, we developed three measures: a *traditional* measure using binary RUCA codes (rural or urban), a *developed land* measure using a continuous variable derived from the percentage of built-up land in each zip code, and a *distance to care* measure in which we calculated actual travel time to care.

The traditional approach involved classifying zip codes as rural or urban variables using RUCA classification for zip codes, which are available in 2-, 4-, or 9-level classifications. The Rural Health Research Center publishes a publicly available crosswalk linking 2004 zip codes to RUCA codes (WWAMI RUCA Rural Health Research Center, n.d.).

The developed land approach involved construction of a rurality variable for each zip code that represented the percentage of developed land in each zip code using data 2011 National Land Cover Database (NLCD) produced by the US Geological Survey (Wickham et al., 2014). The NLCD is a 30-meter spatial resolution raster dataset that categorizes all surface area of the United States into one of 20 different land cover types (such as barren land, evergreen forest or cultivated crops). Four out of these 20 types are regarded as developed, ranging from developed open space to high intensity developed (Homer et al., 2012). The Reclassify tool and the Zonal Statistics tool in ESRI ArcGIS (version 10.6; Redlands, California, USA) were used to classify all land cover types into two distinct categories of land (developed vs. non-developed), and calculate the resultant statistic that represented the percent of developed land area within each zip code.

For the distance to care approach, we initially planned to determine the distance from the geographic center of each child’s zip code to the closest pediatric primary care provider (PCP). However, gathering location data for each pediatric PCP’s office proved to be prohibitively challenging. We determine that multiple types of providers would need to be included, such as pediatrician offices, family practice offices, and free clinics, and we found no address lists containing data where these services were provided. Density data on certain practice types can be obtained through the Area Resources File, but these data are provided only at a county level and would not yield granular data necessary for a meaningful rural analysis (Department of Health and Human Services, 2019). Because our analysis was focused on access to WCVs for the purpose of developmental screenings and may be a proxy of area-level knowledge of pediatric developmental delays, we chose instead to calculate the distance to the closest developmental pediatrician. We used ESRI StreetMap USA data and network routing function in ArcGIS to calculate driving time from the geographic center of each zip code to the closest developmental pediatrician. The geographic center was determined with a population-weighted zip code centroid. To account for our early childhood sample, population weights were developed with population of children under 5 years old in each CBG from 2010 U.S. Census data. Identification of developmental pediatricians was conducted in a manner similar to prior studies, focusing on an extensive internet search verified by a list from the Virginia Department of Health (DeGuzman, Altrui, et al., 2017). The closest five distances were combined using a weighted mean, with the closest distances having the highest weights.

## Data Analysis

Well child visit attendance rate was calculated as a proportion of visits attended between the 12-month and pre-kindergarten visits. Well child attendance rate was treated as a continuous variable and was analyzed using t-tests to compare groups (urban vs. rural). Because each patient visit was associated with a zip code, we selected to define each child’s rurality status using the zip code at the time of the pre-kindergarten visit, although most children in our data set did not move zip codes during the study period. For the traditional approach, zip codes were mapped to binary RUCA codes using publicly-available mapping guidelines (WWAMI RUCA Rural Health Research Center, n.d.). Chi-square tests were used to test for association between categorical variables (e.g. autism diagnosis and urban/rural status). The relationship between continuous variables (distance to care, developed land) and visit attendance rate was analyzed using Spearman correlation and linear regression. Multiple linear regression was used to adjust for payer in analyses of distance to care and developed land.

## Results

The children’s characteristics are presented in Table 1. The 6349 children in the sample were split roughly equally between male and female. Over 75% of the children’s race data was missing. Nearly 82% of the children lived in a ZIP code classified as “urban” by RUCA codes. The majority of children had private insurance (54.1%), with the bulk of the remainder insured by Medicaid (45.8%). Because the dataset included only insurance claims, no self-pay visits were reported.

**Table 1.** Sample Characteristics (n=6349)

| Characteristic | Levels | Frequency | Percent |
| --- | --- | --- | --- |
| Gender | Male | 3217 | 50.67 |
|  | Female | 3122 | 49.17 |
|  | Gender Unknown | 10 | 0.16 |
| Race |  |  |  |
|  | American Indian / Alaska Native | 4 | 0.06 |
|  | Asian | 89 | 1.40 |
|  | African American | 596 | 9.39 |
|  | Hawaiian or Pacific Islander | 5 | 0.08 |
|  | White | 874 | 13.77 |
|  | Other or Unknown or Missing | 4781 | 75.30 |
| Insurance Type |  |  |  |
|  | Medicaid | 2906 | 45.77 |
|  | Private | 3433 | 54.07 |
|  | Missing | 10 | 0.16 |
| ZIP code rurality |  |  |  |
|  | Rural | 1150 | 18.11 |
|  | Urban | 5189 | 81.72 |
|  | Missing | 10 | 0.16 |

Note: Rural/Urban dichotomized using Rural Urban Commuting Area codes

The overall mean WCV attendance rate was 66.7% (range 17% - 83%). Table 2 shows the overall attendance rate by visit for the early childhood WCVs. The visit attendance at the 18-, 24- and 30-month visits (when ASD and developmental screenings are recommended to be performed) are 81.7%, 78.8%, and 26.6% respectively. Table 3 shows the impact of insurance type on WCV rates. The median and mean are both higher for children with private insurance than for those with Medicaid (66.7% and 61.8% vs 50% and 49% respectively, p<0.0001).

**Table 2.** Early Childhood Well Child Visit Attendance Rates (n=6349)

| Well Child Visit | Attendance | Rural (n=1150) | Urban (n=5189) |
| --- | --- | --- | --- |
|  | Frequency (percent) |  |  |
| 12 months | 6349 (100.0) | 1150 (100) | 5189 (100%) |
| 15 months | 4304 (67.8) | 776 (67.5) | 3528 (68.0) |
| 18 months | 5174 (81.5) | 908 (79.0) | 4266 (82.2) |
| 24 months | 4993 (78.6) | 849 (73.8) | 4144 (79.9) |
| 30 months | 1682 (26.5) | 261 (22.7) | 1421 (27.4) |
| 3 years | 4665 (73.5) | 764 (66.4) | 3901 (75.2) |
| 4 years/Pre-K | 6349 (100.0) | 1150 (100.0) | 5189 (100.0) |

\*Note: For children to be included in the dataset they had to have a 12 month Well Child Visit (WCV) and at least one WCV between ages 4-6.

**Table 3.** Impact of insurance type on well child visit attendance rates (n=6339)

| Insurance* | N | Lower Quartile | Median | Upper Quartile | Mean | Maximum |
| --- | --- | --- | --- | --- | --- | --- |
| Private | 3433 | 0.5 | 0.667 | 0.667 | 0.618 | 1 |
| Medicaid | 2906 | 0.333 | 0.5 | 0.667 | 0.493 | 1 |

\* t-test shows a difference between rural and urban zip codes, $p < .0001$ .

### Traditional Rural Analysis

Using RUCA dichotomous codes to classify zip codes as rural or urban, children living in urban areas of Virginia have higher rates of attending early childhood WCVs than their rural counterparts (67% vs. 50% respectively, p<.0001). Table 4 presents full results of the traditional rural analysis. Figure 1 presents box plots of WCV stratified by rurality and insurance payer, to visually identify the relative impact of each of these variables on WCV rates. Within payers, higher attendance rates for urban children can still be seen for those with private insurance; while rates for rural and urban children with Medicaid appear similar.

**Figure 1:** Boxplots of well child visit attendance rates stratified by insurance type and rurality status. Insurance is recorded at the pre-kindergarten visit. On each boxplot, parameters of each quartile are by horizontal lines and the means are marked with circles. On the private insurance boxplots, the 75^th^ percentile and median horizontal lines are sufficiently close as to appear on the same line, around the 65^th^ percentile.

**Table 4.**
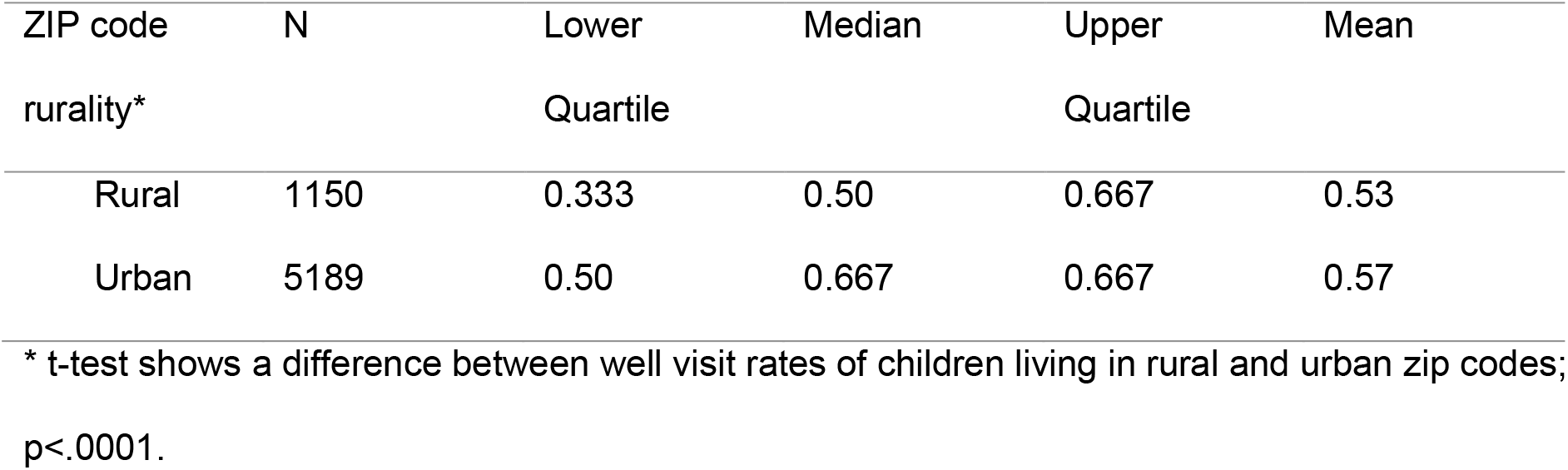
Impact of rurality on well child visits attendance rates using binary classification of Rural Urban Commuting Area Codes (n=6349)

### Developed Land

The developed land variable was developed for 896 Virginia zip codes. Figure 2a shows the state of Virginia with each zip code shaded by its percent of developed land, and 2b shows a histogram of the developed land variable for all Virginia zip codes. The histogram suggests a bimodal distribution, where the majority of zip codes are either highly developed, or less developed, appropriately reflecting Virginia’s mix of highly urban and rural landscapes. Linear regression confirms that the developed land variable identifies a difference in WCVs, with a coefficient estimate of 0.035 indicating that for every increased percentage of developed land, there is a 0.035 increase in WCV attendance (p<0.0001). Applying our findings to an example Virginia zip code suggests that children living in Norfolk, Virginia (zip 23507, 100% developed land) have a 3.8% higher WCV attendance rate than those living in White Post, Virginia (zip code 22663, 8.8% developed land). There is a significant relationship between insurance payer and developed land: children with private insurance live in more urban areas of Virginia (47% developed vs 41% for Medicare, p<.0001). Adjusting for payer, the association between WCV rates and developed land is still significant, but is attenuated at 0.0189.

**Figure 2a:** Virginia, USA. Each zip code is shaded by the percent of developed land in that zip code, using data from the National Land Use Database. Darker shading indicates less developed land.

**Figure 2b:** Histogram showing the percent of Virginia zip codes that fall into categories of developed land percent.

### Distance to Care

Fifty-seven developmental pediatrician locations were identified in Virginia and neighboring states. Figure 3a shows a map of Virginia with zip codes shaded with the resultant weighted mean travel time to reach the closest 5 developmental pediatricians. Spearman correlations show a lack of relationship between travel time and WCV (r= -0.13, p=ns; r=-0.12, p=ns, respectively). The scatterplot in Figure 3b incorporates the children’s’ insurance providers, which shows no demonstrable impact of payer. However, adding payer to the model further weakens the association between WCV visits and distance to care even further.

**Figure 3a:** Virginia, USA. Each zip code is shaded by travel time in minutes from each participants’ home to the closest developmental pediatrician. Location of participants’ homes are estimated using the under 5 years old population-weighted zip code centroid. Closest developmental physician is the weighted mean of the closest five developmental pediatrician addresses. Darker shading indicates a closer distance to care.

**Figure 3b:** Scatterplot. Travel time in minutes between participants’ homes and the closest developmental physician, calculated as described in Figure 3a. Travel time is plotted against the percent of attended well child checks for that participant. Data is stratified by payer.

## Discussion

Our analysis suggests that Virginia rural children attend significantly fewer WCVs during early childhood than their urban counterparts. Our findings are consistent with other studies of early childhood access to care, particularly studies that have found disparities among early childhood for access to specialty care and dental care in rural areas (Martin et al., 2012; McManus et al., 2016). Within this context, our results suggest that during early childhood, rural children are seeing healthcare providers far less frequently than urban children. Because of the emphasis of identifying developmental delays during early childhood, attending fewer primary care visits may make children living in rural Virginia more susceptible to late identification of and delayed treatment for developmental disorders such as ASD.

Our research shows that access to private insurance significantly improves WCV attendance for children, which is similar to earlier findings of lower outpatient visit utilization among Medicaid-enrolled children compared to those with private insurance (Chang et al., 2014). However, the difference in visit attendance may not be directly related to being enrolled in a public insurance program. In Virginia, access to private insurance likely represents other factors that may impact WCV attendance, such as higher family income and parental education, improved access to transportation, and more parental job flexibility compared to children enrolled in Medicaid. In order to develop interventions to improve children’s access to primary care, the separate impact of these factors need further investigation. However, if we assume that insurance type may be a proxy for several social determinants of health, we can view the impact of rurality in this analysis as having a separate and distinct impact from other social determinants.

### Impact of the Coronavirus Disease Pandemic on Well Child Visits

Our data pre-dates the current coronavirus disease pandemic (COVID-19), but the potential impact of COVID-19 on WCV attendance during early childhood needs to be considered. Once lockdown procedures were initiated across the U.S., children’s vaccination rates plummeted, particularly for children over 2 years of age (Santoli et al., 2020). Although data evaluating WCV during this period has not yet been evaluated, it is likely that this abrupt drop is associated with a parallel reduction in WCVs. During COVID-19, many providers transitioned to telemedicine video visits to ensure adequate access (Katzow, Steinway, & Jan, 2020). However, success with conducting problem visits using a telemedicine modality does not imply that all components of a WCV would be successful using this medium. Researchers are investigating methods of developmental disorder screening using telehealth, but parity with an in-person visit has yet to be achieved (Juárez et al., 2018; Narzisi, 2020). Even with demonstrated successful telemedicine screening protocols, rural children are more likely to live in broadband-poor areas, further preventing access to appropriate care (Douthit et al., 2015; Whitacre et al., 2017).

Our data in the context of limited vaccine administration suggests that today’s young rural children are at an even higher risk of not receiving appropriate developmental screening. Nurses caring for young children who live in rural areas need to be aware of these existing and potential disparities. Encouraging parents of rural children with strategies to keep WCV appointments for their children may help avert a cohort of children who experience delayed treatment for ASD. Research needs to evaluate any increases in delays so that public health strategies can be designed quickly to prevent broadening of these disparities.

### Rurality Metrics

Our analysis demonstrates how varying the method of measuring rurality yields different results, and suggest that using a more precise method (such as developed land) may yield more useful information for identifying certain disparities. Two methods of measuring rurality, using RUCA codes and the developed land variable, resulted in a finding a significant impact of rurality on WCV rates. However, using the developed land variable yields potentially more accurate, precise and actionable information. The impact can be seen clearly in the comparison of zip codes in White Post, and Norfolk, Virginia. Using RUCA, the zip codes for these areas Virginia are both considered 1/urban and therefore would have similar expected levels of WCV rates in an analysis using RUCA, regardless of applying a 2-level, 4-level, or 9-level categorization. The developed land variable may result in greater precision for identifying where pre-school children lacking health prevention may live. Use of this variable provides a continuous rather than categorical variable, which may not only be more accurate for some rural health disparities research, but the precision of the variable makes it amendable to prioritization of geographically-targeted area-level interventions. Using distance to care as a rurality variable did not yield a significant result, despite utilizing a network approach, which best approximates real world geographic accessibility. We believe this is largely due to the selection of developmental pediatricians as a proxy for distance to care. Developmental pediatricians are highly clustered in large, densely populated areas, or near a large university (DeGuzman, Altrui, et al., 2017). However, it is important to note that imprecise, population-weighted zip codes as a proxy for address likely affected the accuracy of this metric, as with all of our rurality approaches. Zip codes typically cover far more land area in rural zip codes compared with urban ones, and although we adjusted for a population-weighted centroid, road distribution is also not equally distributed in large area zip codes.

While density and distance to care are difficult to quantify using available data, overall the impact of rurality on access to care may best be embedded in the developed land variable. Preventive healthcare practices are part of the built environment because they require bricks-and-mortar buildings for the physical examination of patients. The physical footprint of each practice is directly built into the developed land variable, but not into the other two approaches.

## Limitations

There are several factors limiting the interpretability of these results. First, our data includes only those children receiving WCVs in Virginia during the study time. It excludes those children who moved in and out of the state, those who may have lived in the state but stopped seeing a clinician for WCVs after birth, and children without any health insurance. Including those children may have changed our results to reflect either a larger or smaller difference between rural and urban children with respect to WCV attendance. Second, we were unsuccessful in conducting a true distance to care analysis. Our efforts were limited by a lack of availability of comprehensive address data for pediatric PCPs. Because these analyses are important to demonstrate an impact of access disparities on early childhood developmental screenings, additional methods should be attempted. Recent technology has improved the ability of public health researchers to gather up large amounts of publicly available data via *web scraping*, the use of computer programs to gather and organize large amounts of data from the World Wide Web (Rennie et al., 2020), which could be attempted to support a more robust analysis, which might show a relationship between distance to PCP as a determinant of WCVs. Finally, our intent was to conduct an exploration of the impact of rurality on WCVs, and understanding how changing rurality variables could affect this. Our study is not meant to be interpreted as a definitive analysis of all socioeconomic factors impacting WCV attendance rates in early childhood, although it is likely that insurance status is a proxy for many of these determinants. Our intent was to continue the discussion of the vulnerabilities of rural children with respect to preventative health care, draw increased attention to the way we analyze and interpret rural disparities research, and recommend a better approach.

## Conclusion

Young children in rural Virginia attend fewer WCVs than their non-rural counterparts, placing them at higher risk for missing vaccinations and developmental disorder screenings. These findings take on additional significance in the context of COVID-19, when rates of vaccinations have plummeted. Future research needs to explore the impacts of COVID-19 on WCV disparities for rural children. As trusted professionals, pediatric nurses should encourage families of young children to continue WCVs when possible, to ensure that missed developmental milestones are identified and addressed in a timely manner.

## Data Availability

Data is available through the Virginia All-Payer Claims Database

